# Ensemble Forecasts of Coronavirus Disease 2019 (COVID-19) in the U.S.

**DOI:** 10.1101/2020.08.19.20177493

**Authors:** Evan L Ray, Nutcha Wattanachit, Jarad Niemi, Abdul Hannan Kanji, Katie House, Estee Y Cramer, Johannes Bracher, Andrew Zheng, Teresa K Yamana, Xinyue Xiong, Spencer Woody, Yuanjia Wang, Lily Wang, Robert L Walraven, Vishal Tomar, Katharine Sherratt, Daniel Sheldon, Robert C Reiner, B. Aditya Prakash, Dave Osthus, Michael Lingzhi Li, Elizabeth C Lee, Ugur Koyluoglu, Pinar Keskinocak, Youyang Gu, Quanquan Gu, Glover E. George, Guido España, Sabrina Corsetti, Jagpreet Chhatwal, Sean Cavany, Hannah Biegel, Michal Ben-Nun, Jo Walker, Rachel Slayton, Velma Lopez, Matthew Biggerstaff, Michael A Johansson, Nicholas G Reich, on behalf of the COVID-19 Forecast Hub Consortium

**Author notes:** corresponding authors: Michael A Johansson, Nicholas G Reich. See full consortium author list in Supplement (on behalf of the COVID-19 Forecast Hub Consortium). The findings and conclusions in this report are those of the authors and do not necessarily represent the views of the Centers for Disease Control and Prevention.

## Abstract

**Background:** The COVID-19 pandemic has driven demand for forecasts to guide policy and planning. Previous research has suggested that combining forecasts from multiple models into a single “ensemble” forecast can increase the robustness of forecasts. Here we evaluate the real-time application of an open, collaborative ensemble to forecast deaths attributable to COVID-19 in the U.S.

**Methods:** Beginning on April 13, 2020, we collected and combined one- to four-week ahead forecasts of cumulative deaths for U.S. jurisdictions in standardized, probabilistic formats to generate real-time, publicly available ensemble forecasts. We evaluated the point prediction accuracy and calibration of these forecasts compared to reported deaths.

**Results:** Analysis of 2,512 ensemble forecasts made April 27 to July 20 with outcomes observed in the weeks ending May 23 through July 25, 2020 revealed precise short-term forecasts, with accuracy deteriorating at longer prediction horizons of up to four weeks. At all prediction horizons, the prediction intervals were well calibrated with 92-96% of observations falling within the rounded 95% prediction intervals.

**Conclusions:** This analysis demonstrates that real-time, publicly available ensemble forecasts issued in April-July 2020 provided robust short-term predictions of reported COVID-19 deaths in the United States. With the ongoing need for forecasts of impacts and resource needs for the COVID-19 response, the results underscore the importance of combining multiple probabilistic models and assessing forecast skill at different prediction horizons. Careful development, assessment, and communication of ensemble forecasts can provide reliable insight to public health decision makers.

## Introduction

The outbreak of Coronavirus Disease 2019 (COVID-19) in Wuhan, China, in late December 2019 quickly spread around the world, resulting in formal recognition of COVID-19 as a global threat by the World Health Organization on January 30, 2020 (Promed 2019; World Health Organization 2020). Subsequent rapid, global spread of SARS-CoV-2, the virus that causes COVID-19, drove an urgent need for forecasts of the timing and intensity of future transmission and the locations with the highest risk of spread to inform risk assessment and planning.

Multiple studies of epidemic forecasting have shown that ensemble forecasts, which incorporate multiple model predictions into a combined forecast, consistently perform well and often outperform most if not all individual models (Viboud et al. 2018; Johansson et al. 2019; McGowan et al. 2019; Reich, Brooks, et al. 2019). Ensemble approaches are in widespread use in other fields such as economics (Timmermann 2006; Busetti 2014) and weather forecasting (Leutbecher and Palmer 2008). Ensemble models can distill information across multiple forecasts and are a robust option for decision making and policy planning, especially in situations where extensive historical data on individual model performance are not available.

Here, we summarize a collaborative effort between the U.S. Centers for Disease Control and Prevention (CDC), 21 largely academic research groups, five private industry groups, and two government-affiliated groups. Starting in April 2020, this consortium, called the COVID-19 Forecast Hub (https://covid19forecasthub.org), developed shared forecasting targets and data formats, then constructed, evaluated, and communicated the results of ensemble forecasts for U.S. deaths attributable to COVID-19. Deaths due to COVID-19 are a proximate indicator of burden on health care systems and a critical measure of health impact.

## Methods

Beginning on April 13, 2020 and every Monday thereafter, we collected probabilistic one-, two-, three-, and four-week ahead forecasts of the total number of deaths due to COVID-19 that would be reported by the Center for Systems Science and Engineering (CSSE) at Johns Hopkins University (Dong, Du, and Gardner 2020) by the Saturday of each week for U.S. states and territories and the United States overall. Prediction intervals (e.g., 95% or 50%) characterize uncertainty which point forecasts are unable to characterize. Thus, each participating team provided the median of the predictive distribution and 11 prediction intervals ranging from a 10% prediction interval to a 98% prediction interval. Participating groups were able to use the methods and data sources they deemed appropriate to generate the forecasts (Reich et al. 2020).

On April 27, after two weeks of consistent submissions, we began aggregating individual forecasts of cumulative deaths to construct a weekly ensemble forecast (hereafter referred to as “ensemble”). To be eligible for inclusion in the ensemble, a model had to submit a valid forecast for all four week-ahead horizons (details on validation in Supplemental Figure 1). The total number of individual models included in the ensemble forecasts ranged from six (April 27) to 20 (June 15 and 29). Ensemble forecasts for specific locations often included fewer models because some models did not include forecasts for all locations. The ensemble forecast was constructed as an equally-weighted average of forecasts from all eligible models. Specifically, the endpoints of each prediction interval in the ensemble forecast were calculated as the average of the corresponding quantiles across all individual model forecasts (Busetti 2014). Ensemble forecasts were constructed by researchers at the University of Massachusetts Amherst and posted publicly by CDC each week.

**Figure 1:**
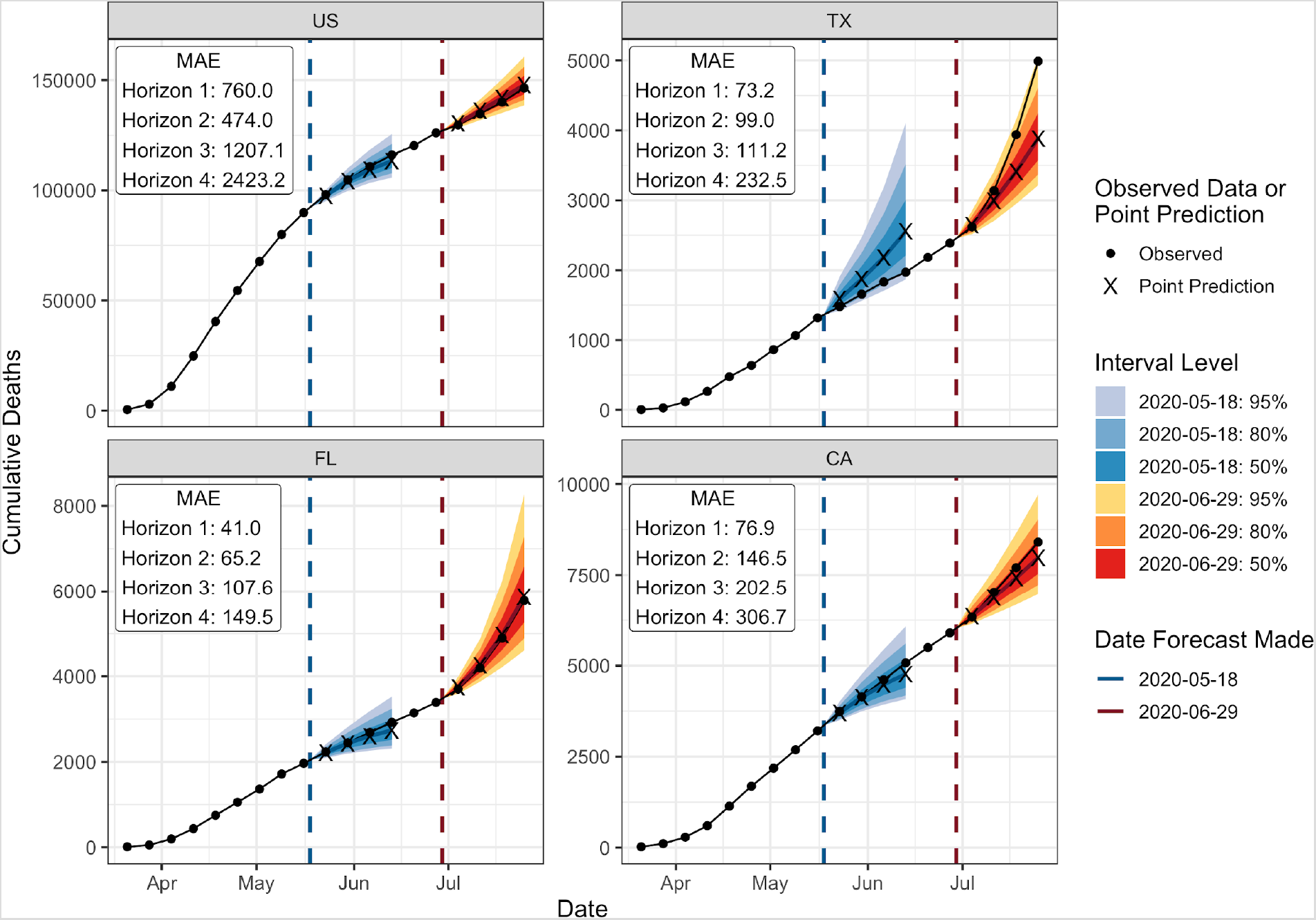
Reported deaths and example forecasts. Reported cumulative deaths due to Coronavirus Disease 2019 (COVID-19) from March 21 to July 25 as of July 26, 2020 (black points) for the United States (US) and the three states with highest reported deaths in the week ending July 25 (Texas [TX], Florida [FL] and California [CA]). Forecasts are one- through four-week ahead predicted medians and 50%, 80%, and 95% prediction intervals from the ensemble forecasts created on May 18 and June 29. Forecasts from these two dates are shown as examples. Ensemble forecasts were made every week starting April 27, 2020 (not shown, available at (Reich et al. 2020)). The mean absolute error (MAE) is the mean of the difference between the forecasted and observed values across all forecasts at each horizon (one- to four-weeks ahead) for which the target was observed as of July 26.

We evaluated ensemble forecasts of cumulative deaths in the United States compared to reported deaths for only weeks which had previous forecasts at all four prediction horizons (one- to four-weeks ahead). For example, we did not evaluate forecasts for the week ending May 2, because only a one-week ahead ensemble forecast was available, making comparison across horizons impossible. Overall, we compared forecasts across nine weeks of observations, from the week ending May 23 through the week ending July 25, 2020. We evaluated the performance of ensemble forecasts across all locations and weeks where at least two individual models contributed to the ensemble; this included 2,512 of the 2,561 possible forecasts during the evaluation timeframe for different locations and forecast horizons. Reported cumulative death data for this evaluation were downloaded from the CSSE repository on July 26, 2020.

We evaluated the error of the ensemble with mean absolute error (MAE), the mean difference between the forecasted and observed values, and calibration with prediction interval coverage. Prediction interval coverage was calculated by determining the frequency with which the prediction interval contained the eventually observed outcome. In a model that accurately characterizes uncertainty, the prediction interval level will correspond closely to the frequency of eventually observed outcomes that fall within that prediction interval. For example, eventually observed values should be within the 95% prediction interval approximately 95% of the time. Because the ensemble prediction intervals were calculated by averaging individual models, the ensemble prediction intervals typically were not whole numbers and almost never included predictions of no new deaths in a given week. We therefore assessed interval coverage for the original ensemble and for the rounded ensemble. We rounded the interval endpoints to a whole number conservatively, rounding the lower limits of prediction intervals down and the upper limits of prediction intervals up.

The forecast data and submission instructions are available in a public GitHub repository (Reich et al. 2020). Analyses were performed in R (https://www.R-project.org/) and all code used to construct the ensemble forecasts and reproduce the analyses in this manuscript is available in a separate public GitHub repository (E. Ray 2020).

## Results

Comparing forecasts to the reported death data for the weeks ending May 23 through July 25, 2020, the ensemble point forecasts were accurate and precise at short-term prediction horizons, with a general increase in error at longer horizons (Figure 1, Supplemental Figure 2, Supplemental Table 1). Specifically, on average across all locations, the mean absolute error (MAE) of four-week ahead point predictions was more than three times the MAE of one-week ahead predictions. For example, the national-scale one-week ahead MAE indicated an average difference of 760 deaths from the eventually observed values, while the four-week ahead forecasts had a MAE of about 2,400 deaths (Figure 1).

The ensemble forecasts were also well calibrated, with prediction intervals that covered the observed data with the expected frequency (Table 1, Supplemental Figure 3). The 50% prediction intervals were conservative; they captured 50-55% of observations for all forecast horizons with the original forecasts, and 57-65% with the conservatively rounded, whole number forecasts. In contrast, the original 95% prediction intervals captured only 87-90% of observations. After rounding, the 95% prediction intervals were better calibrated, with coverage rates of 92-96%. Rounding had a particularly clear impact on forecasts for weeks with no reported deaths as many ensemble forecasts had lower prediction bounds just above zero due to one or more individual forecasts being greater than zero. For example, rounding changed the 95% coverage for 120 forecasts and 99 of those were for weeks with no new deaths. While the forecasts were less precise at longer horizons (greater MAE), they remained calibrated.

**Table 1:** Observed prediction interval coverage for deaths reported May 23 through July 25, 2020 indicating calibration for ensemble forecasts of cumulative deaths in locations with at least two valid individual forecasts. The top section summarizes the original ensemble forecasts, and the bottom section summarizes the ensemble forecasts after rounding the lower limits of prediction intervals down and the upper limits of prediction intervals up.

|  |  | Observed Prediction Interval Coverage |  |  |  |
| --- | --- | --- | --- | --- | --- |
|  |  | Forecast Horizon (weeks ahead) |  |  |  |
|  | Measure | 1 | 2 | 3 | 4 |
| Original Ensemble Forecasts | 50% Coverage | 53.3% | 50.5% | 51.6% | 54.6% |
|  | 95% Coverage | 87.3% | 87.0% | 89.2% | 90.1% |
| Rounded Ensemble Forecasts | 50% Coverage | 65.1% | 59.9% | 57.9% | 57.7% |
|  | 95% Coverage | 95.6% | 93.2% | 93.8% | 92.8% |

## Discussion

The spread of COVID-19 has driven a continually adapting global response. Substantial uncertainty persists about the disease’s trajectory, and robust forecasts of COVID-19 activity can help inform decision making in the face of this uncertainty. The COVID-19 Forecast Hub was created to rapidly collect, aggregate, ensemble, and evaluate forecasts in real-time. The resulting forecasts were published on the CDC website starting on April 27, 2020, (Centers for Disease Control and Prevention 2020) and were used to develop summary messages about the trajectory of the outbreak in the United States. These early results indicate that ensemble forecasts at the national and state-level were accurate, with mean absolute errors well below the maximum number of new deaths reported per week, and well-calibrated, with reasonable prediction interval coverage, especially when evaluated using rounded prediction intervals. These forecasts can be used as situational awareness and planning tools to inform a variety of planning and response decisions such as the implementation of mitigation strategies, the distribution of resources, or vaccine trial site selection (Dean et al. 2020; Wallinga, van Boven, and Lipsitch 2010; Lipsitch et al. 2011).

It is critical that forecasts used to inform public health decisions accurately characterize their uncertainty, and these ensemble forecasts achieved that goal. The ensemble forecasts were conservative for more central prediction intervals such as the 50% prediction interval which captured 50-60% of observations, but were well calibrated at the more important, extreme intervals after rounding. In outbreak forecasting, extreme errors can lead to suboptimal decision-making, such as unexpected shortages in or oversupply of resources. A well-calibrated forecast reduces this risk. For example, only 1% of outcomes should exceed the 99% prediction quantile, and this occurred for 1.4% of forecasts made by the COVID-19 Forecast Hub ensemble.

Although the ensemble performed well, there are many avenues for further improvement. The ensemble forecast weighted the models equally, but weighting based on historical performance may improve forecast skill (McAndrew and Reich 2019; Reich, McGowan, et al. 2019; Viboud et al. 2018; E. L. Ray and Reich 2018; Yamana, Kandula, and Shaman 2016; Reis et al. 2019; Brooks et al. 2018). However, any weighting method will need to be dynamic and allow for the incorporation of new models, potential changes in existing models, and the different sets of locations forecasted by each model.

The ensemble forecasts evaluated here only looked four weeks into the future. Although probabilistic forecasts at longer horizons may characterize uncertainty correctly, in this study we observed significant widening of prediction intervals and degrading precision of point forecasts, even at horizons of three- to four-weeks ahead. This raises concerns about the reliability of forecasts at longer horizons of weeks to months. In addition to current transmission dynamics and mitigation measures, long-term forecasts must also predict changes in mitigation and human behavior to inform projections. Uncertainties about data, behavior, and mitigation measures compound at longer horizons and may further reduce precision and accuracy.

This analysis indicates that ensemble forecasts of cumulative mortality generated in real-time at the national and state level during the first six months of the COVID-19 outbreak in the United States were accurate and well-calibrated. Given this and the previous performance of probabilistic ensemble forecasts for multiple infectious diseases, we encourage decision makers to consider the use of multiple models and ensemble forecasts rather than single model forecasts for future risk assessment and planning needs.

## Data Availability

All data and code referred to in the manuscript are publicly available.

https://github.com/reichlab/covid19-forecast-hub/

https://github.com/reichlab/covidEnsembles

https://zoltardata.com/project/44

## Supplement

**Supplemental Figure 1:**
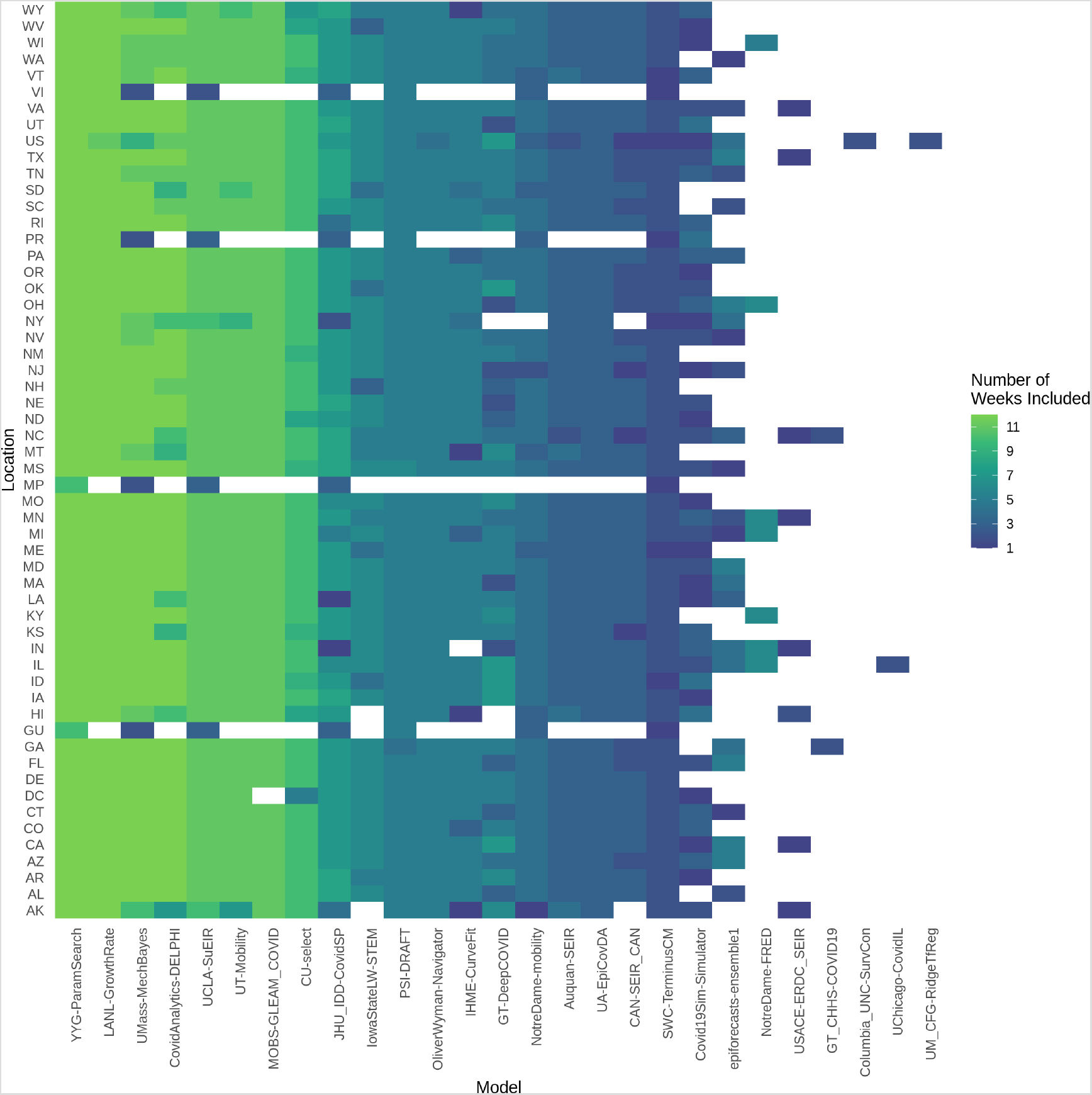
The number of times each contributing model was in the ensemble forecast for each location. An empty cell indicates that the model was not included in a forecast for the given location. The number of individual models included varied because of differences in number of submissions, locations included in submissions, and the following criteria for individual forecasts: (1) a forecast had to include all four week-ahead horizons, (2) the one week ahead forecast for cumulative deaths should not assign probability more than 0.1 to a reduction in cumulative deaths relative to already reported deaths, and (3) at each quantile level, predictions should be non-decreasing over the four prediction horizons. Abbreviations for each location are shown in Supplemental Table 1.

**Supplemental Figure 2:**
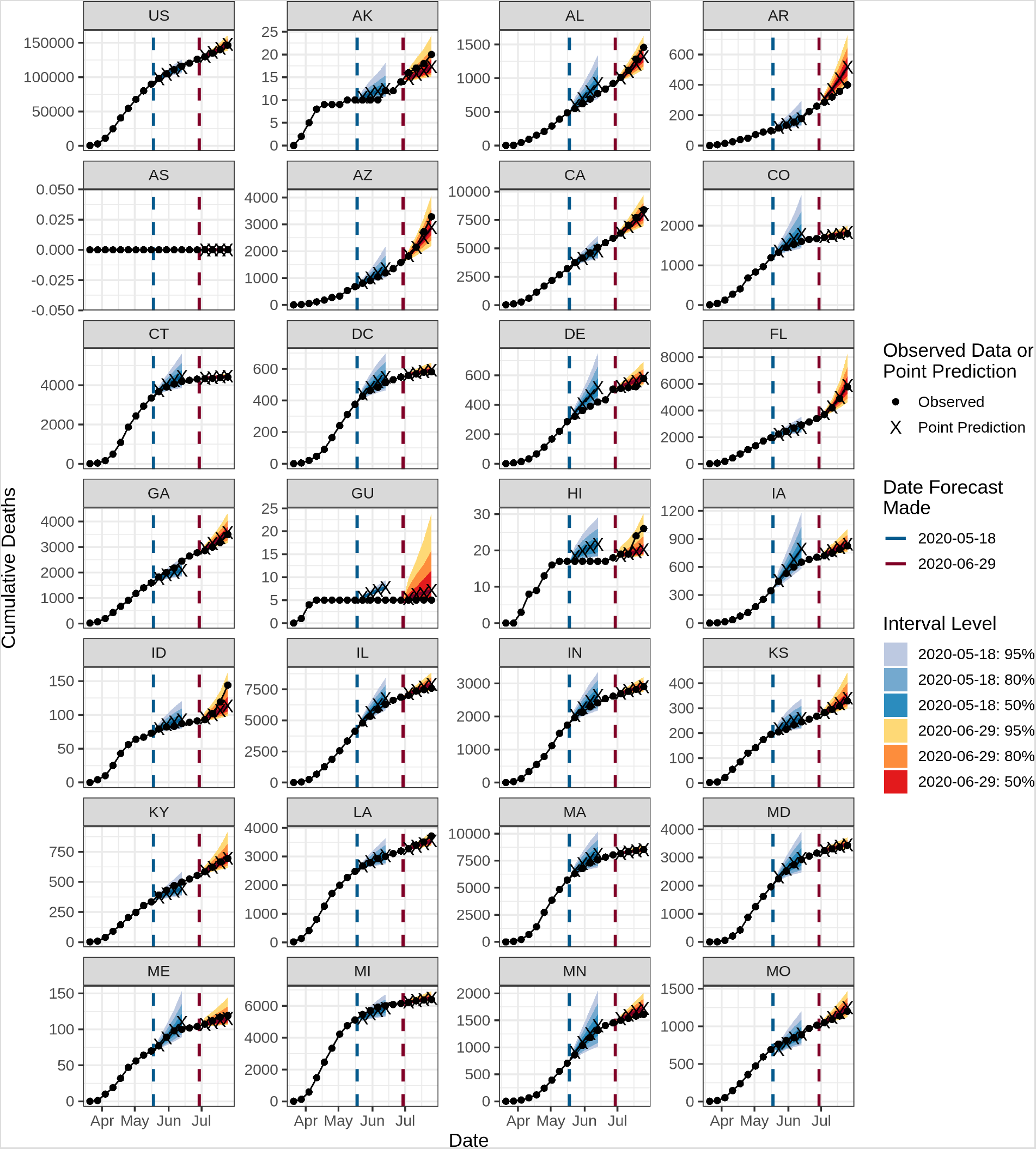
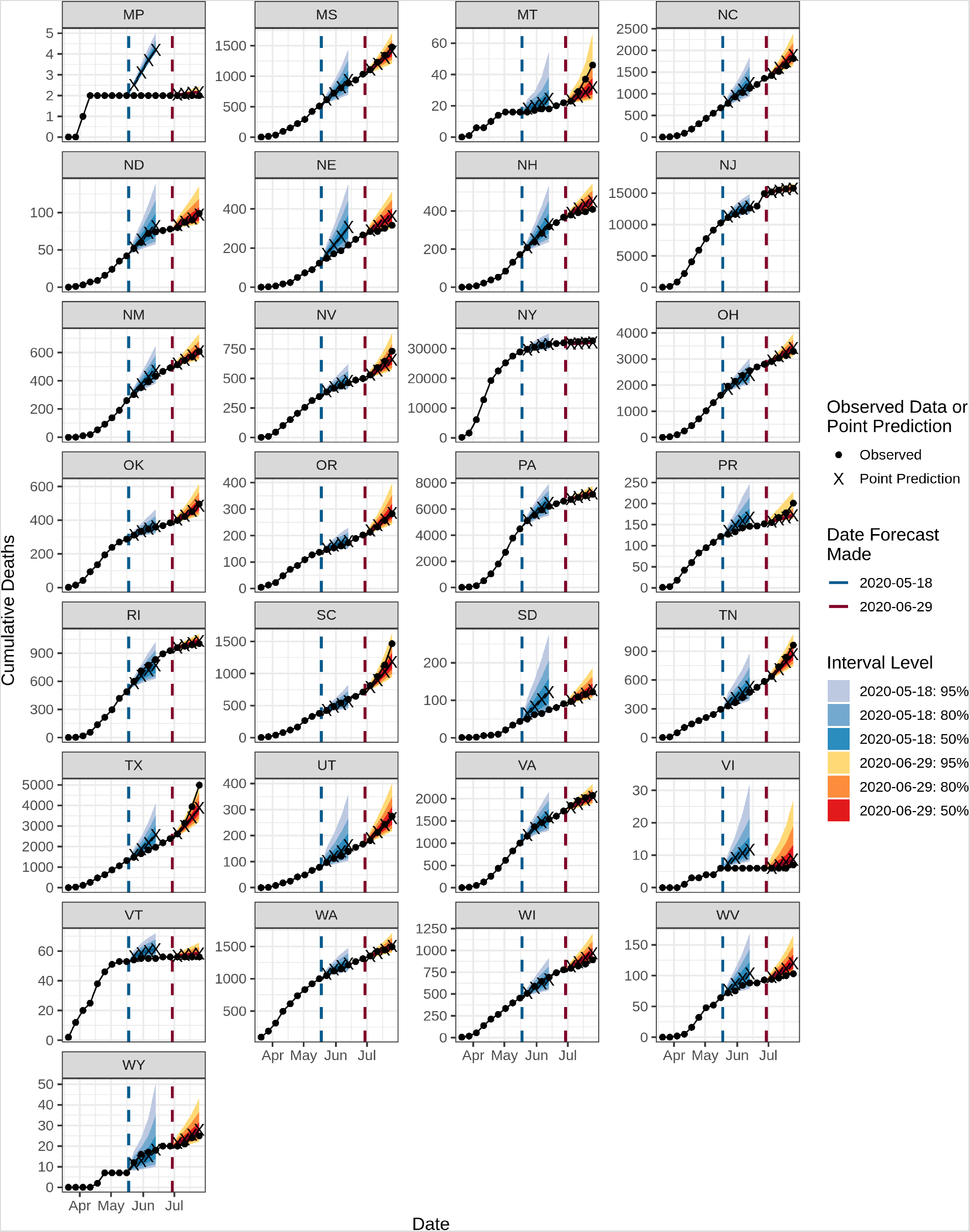
Reported cumulative deaths due to COVID-19 for the US and states and territories from March 21 to July 25 as of July 26, 2020. Forecasts are 1- through 4-week ahead predicted medians and 50%, 80%, and 95% prediction intervals from the ensemble forecasts created on May 18 and June 29. Forecasts from these two dates are shown as examples; ensemble forecasts were made every week starting April 27, 2020 (not shown, available at (Reich et al. 2020)). Abbreviations for each location are shown in Supplemental Table 1.

**Supplemental Table 1:** MAE for cumulative deaths reported May 23 through July 25, the maximum number of deaths reported per week up through the week ending July 25, and the cumulative deaths reported through the week ending July 25. All reported data were collected on July 26, 2020. Locations are sorted in decreasing order of cumulative deaths. The top row summarizes MAE for all locations other than American Samoa, for which 0 deaths were observed during the evaluation period.

|  |  | Mean absolute error (MAE) |  |  |  |  |  |
| --- | --- | --- | --- | --- | --- | --- | --- |
| Location abbreviation | Location name | 1-week horizon | 2-week horizon | 3-week horizon | 4-week horizon | Maximum weekly deaths | Cumulative Deaths |
| - | All | 49.1 | 68.4 | 112.1 | 175.4 |  |  |
| US | United States | 760.0 | 474.0 | 1,207.1 | 2,423.2 | 15,665 | 146,460 |
| NY | New York | 512.3 | 500.4 | 601.7 | 694.4 | 6,667 | 32,608 |
| NJ | New Jersey | 293.1 | 560.6 | 818.0 | 830.3 | 2,024 | 15,776 |
| MA | Massachusetts | 65.8 | 101.0 | 177.3 | 448.4 | 1,326 | 8,510 |
| CA | California | 76.9 | 146.5 | 202.5 | 306.7 | 706 | 8,408 |
| IL | Illinois | 104.6 | 209.5 | 263.4 | 272.6 | 790 | 7,589 |
| PE | Pennsylvania | 36.2 | 135.3 | 281.0 | 485.5 | 1,084 | 7,124 |
| MI | Michigan | 107.5 | 132.0 | 137.1 | 149.3 | 973 | 6,400 |
| FL | Florida | 41.0 | 65.2 | 107.6 | 149.5 | 882 | 5,777 |
| TX | Texas | 73.2 | 99.0 | 111.2 | 232.5 | 1,051 | 4,990 |
| CT | Connecticut | 26.3 | 59.2 | 94.5 | 202.4 | 779 | 4,413 |
| LA | Louisiana | 11.8 | 19.5 | 48.0 | 98.9 | 461 | 3,715 |
| GA | Georgia | 51.1 | 110.2 | 161.5 | 184.7 | 325 | 3,494 |
| MD | Maryland | 35.2 | 89.9 | 150.7 | 255.0 | 454 | 3,433 |
| OH | Ohio | 44.7 | 92.5 | 136.0 | 203.4 | 346 | 3,297 |
| AZ | Arizona | 31.4 | 52.1 | 97.7 | 133.7 | 579 | 3,286 |
| IN | Indiana | 24.5 | 66.5 | 124.2 | 232.2 | 375 | 2,895 |
| Location abbreviation | Location name | 1-week horizon | 2-week horizon | 3-week horizon | 4-week horizon | Maximum weekly deaths | Cumulative Deaths |
| VI | Virginia | 47.3 | 95.4 | 138.1 | 187.4 | 211 | 2,075 |
| NC | North Carolina | 32.1 | 73.9 | 130.4 | 193.5 | 163 | 1,811 |
| CO | Colorado | 18.1 | 53.6 | 80.3 | 140.2 | 273 | 1,794 |
| MN | Minnesota | 30.1 | 94.3 | 186.1 | 305.1 | 175 | 1,611 |
| WA | Washington | 13 | 17.3 | 24.2 | 27.3 | 180 | 1,494 |
| MS | Mississippi | 18.2 | 26.3 | 62.5 | 92.3 | 132 | 1,478 |
| SC | South Carolina | 14.4 | 25.9 | 45.8 | 67.9 | 330 | 1,465 |
| AL | Alabama | 16.2 | 26.5 | 50.9 | 78.6 | 172 | 1,456 |
| MO | Missouri | 26 | 41.2 | 56.8 | 72.9 | 122 | 1,200 |
| RI | Rhode Island | 13 | 25.4 | 34.9 | 79.8 | 122 | 1,002 |
| TN | Tennessee | 12.3 | 17.3 | 20.3 | 42.4 | 126 | 964 |
| WI | Wisconsin | 16.4 | 37.9 | 64.0 | 101.4 | 83 | 891 |
| IA | Iowa | 15.6 | 44.1 | 93.8 | 201.6 | 100 | 826 |
| NV | Nevada | 9.6 | 19.3 | 22.9 | 28.1 | 86 | 732 |
| KY | Kentucky | 8.8 | 17.4 | 32.2 | 60.6 | 61 | 696 |
| NM | New Mexico | 5.9 | 11.5 | 18.1 | 26.6 | 68 | 607 |
| DC | Washington DC | 7.9 | 19.5 | 32.6 | 47.8 | 75 | 581 |
| DE | Delaware | 15.7 | 26.5 | 35.0 | 38.9 | 73 | 579 |
| OK | Oklahoma | 4.3 | 11.0 | 20.0 | 27.2 | 58 | 496 |
| NH | New Hampshire | 6.5 | 10.0 | 18.2 | 34.1 | 47 | 409 |
| Location abbreviation | Location name | 1-week horizon | 2-week horizon | 3-week horizon | 4-week horizon | Maximum weekly deaths | Cumulative Deaths |
| AR | Arkansas | 7.2 | 16.6 | 18.2 | 15.9 | 47 | 399 |
| KS | Kansas | 3.6 | 11.6 | 23.3 | 55 | 35 | 329 |
| NE | Nebraska | 10.7 | 24.8 | 47.6 | 105.1 | 33 | 316 |
| OR | Oregon | 3.8 | 7.0 | 8.6 | 10.6 | 26 | 282 |
| UT | Utah | 6.9 | 11.5 | 19.2 | 27.8 | 31 | 274 |
| PR | Puerto Rico | 4.9 | 7.9 | 13.0 | 16.8 | 24 | 201 |
| ID | Idaho | 2.1 | 3.4 | 5.1 | 9.2 | 25 | 144 |
| SD | South Dakota | 4.8 | 6.5 | 12.2 | 21.1 | 13 | 122 |
| ME | Maine | 2.1 | 4.9 | 8.3 | 14.4 | 15 | 119 |
| WV | West Virginia | 3.3 | 6.0 | 10.7 | 16.9 | 16 | 103 |
| ND | North Dakota | 3.6 | 6.7 | 11.4 | 17.7 | 12 | 99 |
| VT | Vermont | 1.1 | 2.7 | 6.0 | 10.7 | 13 | 56 |
| MO | Montana | 1.2 | 1.4 | 2.7 | 5.2 | 9 | 46 |
| HI | Hawaii | 0.7 | 1.6 | 3.3 | 4.2 | 5 | 26 |
| WY | Wyoming | 1.0 | 2.1 | 2.7 | 3.6 | 5 | 25 |
| AK | Alaska | 1.0 | 1.4 | 1.7 | 2.4 | 3 | 20 |
| VI | Virgin Islands | 0.6 | 1.4 | 2.0 | 2.6 | 2 | 7 |
| GU | Guam | 0.3 | 0.6 | 0.8 | 1.1 | 3 | 5 |
| MP | Northern Mariana Islands | 2.4 | 3.0 | 3.8 | 4.8 | 1 | 2 |
| AS | American Samoa | 0.0 | 0.0 | 0.0 | 0.0 | 0 | 0 |

**Supplemental Figure 3:**
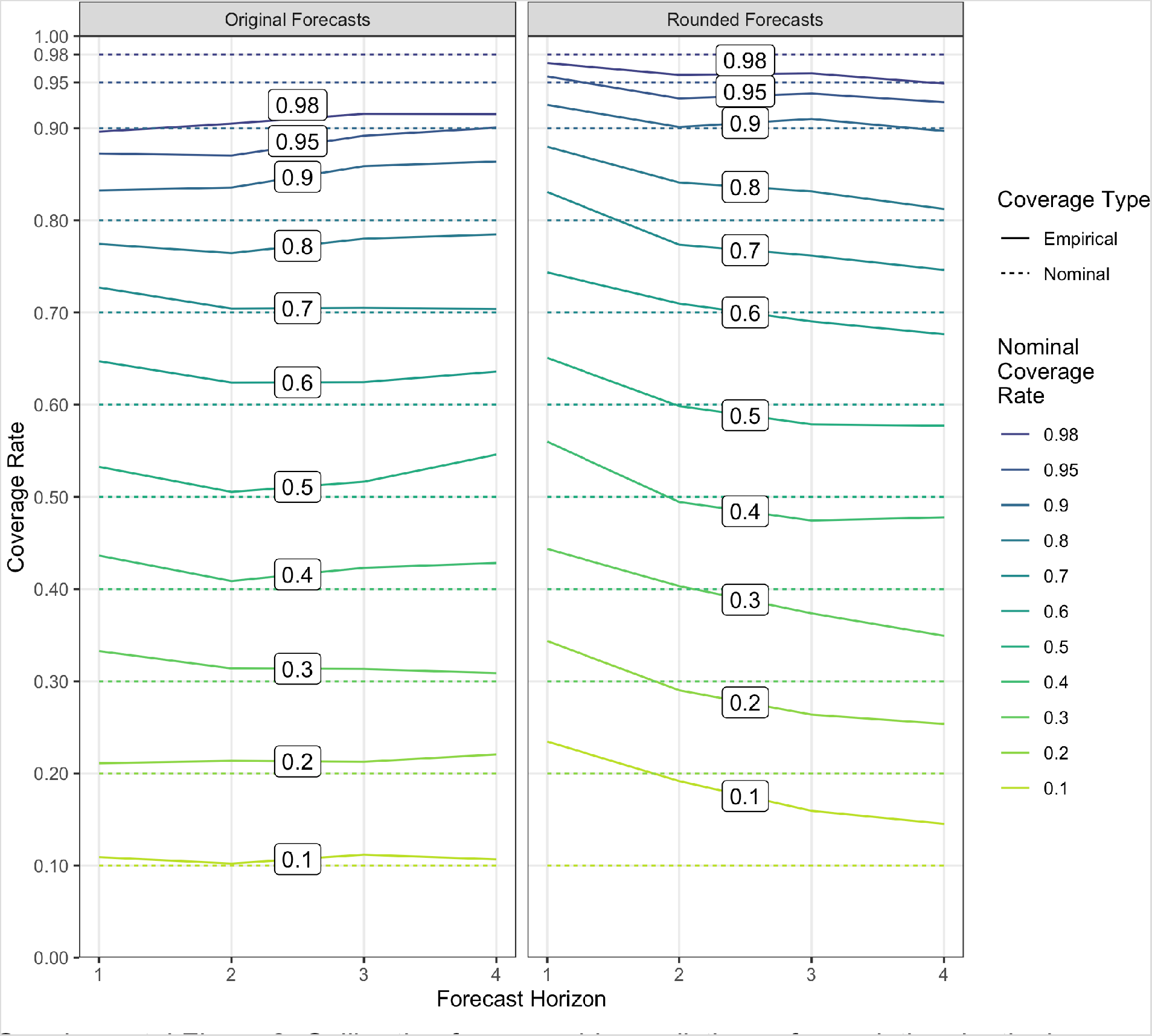
Calibration for ensemble predictions of cumulative deaths in locations with at least two valid individual models submitted. We report calibration of prediction intervals from forecasts for weeks ending May 23 to July 25, 2020, the set of observed weeks with previous forecasts at all four time horizons at the time of writing. Lines are labeled with the nominal coverage rate; a well calibrated forecast will have empirical coverage rate equal to the nominal coverage rate.

## Supplement: COVID-19 Forecast Hub Consortium complete authorship, funding and acknowledgments list

Team name: COVID-19 Forecast Hub

Named Authors: Evan L Ray^1^, Nicholas G Reich^1^, Estee Cramer^1^, Nutcha Wattanachit^1^, Abdul Hannn Kanji^1^, Katie House^1^, Jarad Niemi^2^, Johannes Bracher^3^

Named authors institution:^1^ University of Massachusetts Amherst, School of Public Health and Health Sciences,^2^ Iowa State University, Department of Statistics,^3^ Econometrics and Statistics, Karlsruhe Institute of Technology and Computational Statistics Group and Heidelberg Institute for Theoretical Studies, Germany

Other team authors:

Department of Biostatistics and Epidemiology, University of Massachusetts Amherst: Martha Zorn, Yijin Wang, Matthew Cornell

Department of Biostatistics and Epidemiology, University of Massachusetts Amherst: Khoa Le, Abdul Hannan Kanji, Katie House

Computational Statistics Group, Heidelberg Institute for Theoretical Studies, and Institute for Stochastics, Karlsruhe Institute of Technology, Germany: Tilmann Gneiting

London School of Hygiene and Tropical Medicine: Sebastian Funk

Department of Statistics and Machine Learning Department, Carnegie Mellon University: Ryan J.Tibshirani

Institute of Mathematical Statistics and Actuarial Science, University of Bern: Anja Mühlemann

IQT Labs: Andrea Brennan

Funding: ELR, MZ, YW, NGR, and EC were supported by the US Centers for Disease Control and Prevention (U01IP001122). NGR, NW, and MC were supported by the National Institutes for General Medical Sciences (R35GM119582). RJT was supported by the US Centers for Disease Control and Prevention (U01IP001121). JB was supported by the Helmholtz Foundation via the SIMCARD Information & Data Science Pilot Project. TG is grateful for support by the Klaus Tschira Foundation. AM gratefully acknowledges financial support from the Swiss National Science Foundation. The content is solely the responsibility of the authors and does not necessarily represent the official views of CDC or NIGMS.

Team name: MIT CovidAnalytics

Named Author: Michael Lingzhi Li, MBAn

Named author institution: Operations Research Center, Massachusetts Institute of Technology

Other team authors:

Operations Research Center, Massachusetts Institute of Technology: Hamza Tazi Bouardi, Omar Skali Lami

Sloan School of Management, Massachusetts Institute of Technology: Dimitris Bertsimas, Nikolaos K Trichakis

Brown University: Thomas A Trikalinos

Team name: DeepCOVID

Named Author: B. Aditya Prakash, PhD

Named author institution: College of Computing, Georgia Institute of Technology

Other team authors:

College of Computing, Georgia Institute of Technology: Alexander Rodriguez, Jiaming Cui, Anika Tabassum, Jiajia Xie

Department of Computer Science, University of Iowa: Bijaya Adhikari

Department of Computer Science, University of Illinois Urbana Champaign: Jimeng Sun IQVIA, Cambridge MA USA: Cheng Qian, Cao Xiao

Funding: BAP, AR, JC, AT and JX were partially supported by the National Science Foundation (Expeditions CCF-1918770, CAREER IIS-1750407, RAPID IIS-2027862, Medium IIS-1955883, NRT DGE-1545362), funds from Georgia Tech Research Institute (GTRI) and funds/computing resources from Georgia Tech. The content is solely the responsibility of the authors and does not necessarily represent the views of the funding agencies, Georgia Tech, GTRI, University of Iowa, UIUC or IQVIA.

Team name: University of Michigan

Named Author: Sabrina Corsetti

Named author institution: Physics Department, University of Michigan

Other team authors:

Physics Department, University of Michigan: Ella McCauley

Physics Department, University of Michigan: Tom Schwarz, PhD

Funding: SC and EM, as well as computing resources, were funded by the University of Michigan. The content is solely the responsibility of the authors and does not necessarily represent the views of the University of Michigan.

Team name: Georgia Tech Center for Health and Humanitarian Systems

Named Author: Pinar Keskinocak, PhD

Named author institution: H. Milton Stewart School of Industrial and Systems Engineering, Georgia Institute of Technology

Other team authors:

H. Milton Stewart School of Industrial and Systems Engineering, Georgia Institute of Technology: Buse Eylul Oruc, Arden Baxter, John Asplund, Nicoleta Serban

Funding: PK, BEO, AB, JA, and NS were supported in part by the William W. George and the Virginia C. and Joseph C. Mello endowments at Georgia Tech, the NSF grant MRI 1828187, and research cyberinfrastructure resources and services provided by the Partnership for an Advanced Computing Environment (PACE) at Georgia Tech.

Team name: UCLA Statistical Machine Learning Lab Named Author: Quanquan Gu, PhD

Named author institution: Department of Computer Science, University of California, Los Angeles

Other team authors:

Department of Computer Science, University of California, Los Angeles: Difan Zou, Lingxiao Wang, Pan Xu, Jinghui Chen, Weitong Zhang

Team name: Youyang Gu

Named Author: Youyang Gu, M.Eng Named author institution: None Other team authors: None

Team name: JHU Infectious Disease Dynamics Working Group

Named Author: Elizabeth C Lee, PhD

Named author institution: Department of Epidemiology, Johns Hopkins Bloomberg School of Public Health

Other team authors:

Department of Epidemiology, Johns Hopkins Bloomberg School of Public Health: Justin Lessler, Joshua Kaminsky, Javier Perez-Saez, Juan Dent Hulse, Kyra H Grantz, Stephen A Lauer, Hannah R Meredith

Department of International Health, Johns Hopkins Bloomberg School of Public Health: Shaun A Truelove

International Vaccine Access Center, Johns Hopkins Bloomberg School of Public Health: Shaun A Truelove

Laboratory of Ecohydrology, School of Architecture, Civil and Environmental Engineering, École Polytechnique Fédérale de Lausanne, Lausanne, Switzerland: Joseph C Lemaitre Department of Internal Medicine, Division of Epidemiology, University of Utah: Lindsay T Keegan

Unaffiliated: Kathryn Kaminsky, Josh Wills, Sam Shah

Funding and acknowledgments: ECL, HRM, JCL, JDH, JK, JL, JPS, KHG, LTK, SAL, and SAT were supported by the State of California. ECL, HCL, JK, JL, JPS, LTK, SAL, and SAT were supported by the U.S. Department of Health and Human Services and the U.S. Department of Homeland Security. This work was additionally supported by the Office of the Dean at the Johns Hopkins Bloomberg School of Public Health, the Johns Hopkins Health System, and with computing service credits from Amazon Web Services. These thoughts and opinions are our own and do not represent the views of the U.S. federal government.

Team name: Robert Walraven

Named Author: Robert L Walraven, PhD Named author institution: None

Team name: Auquan

Named Author: Vishal Tomar, MS Named author institution: Auquan Ltd Other team authors:

Auquan Ltd: Chandini Jain

Team name: Columbia_UNC-SurvCon

Named Author: Yuanjia Wang, PhD

Named author institution: Department of Biostatistics, Mailman School of Public Health, Columbia University

Other team authors:

Columbia University: Qinxia Wang, Shanghong Xie

Department of Biostatistics, Gillings School of Public Health, University of North Carolina at Chapel Hill: Donglin Zeng

Funding: YW and DZ are supported by NIH grant GM124104

Team name: Predictive Science Inc.

Named Author: Michal Ben-Nun, PhD

Named author institution: Predictive Science Inc.

Other team authors:

Predictive Science Inc.: Pete Riley, James Turtle

School of Public Health, Imperial College London, UK: Steven Riley

Funding: This work was supported by the Defense Threat Reduction Agency (Award Number: HDTRA1-19-D-0007) and by the National Science Foundation (Award Number: 2031536).

Team name: University of Texas at Austin COVID-19 Modeling Consortium (UT) Named Author: Spencer Woody, PhD

Named author institution: The University of Texas at Austin, Department of Integrative Biology

Other team authors:

The University of Texas at Austin, Department of Statistics and Data Sciences: Mauricio Garcia Tec

Texas Advanced Computing Center: Maytal Dahan, Kelly Gaither

University of Texas, Department of Integrative Biology: Lauren Ancel Meyers, Spencer J Fox

University of Texas McCombs School of Business: James G. Scott

Santa Fe Institute: Michael Lachmann

Team name: University of Massachusetts

Named Author: Daniel Sheldon, PhD

Named author institution: University of Massachusetts Amherst, College of Information and Computer Sciences

Other team authors:

University of Massachusetts Amherst, School of Public Health and Health Sciences: Graham Casey Gibson, Nicholas G. Reich

Team name: University of Arizona - EpiCovDA

Named Author: Hannah Biegel, MSc

Named author institution: University of Arizona, Department of Mathematics Other team authors:

University of Arizona, Department of Mathematics: Joceline Lega

Team name: LANL-GrowthRate

Named Author: Dave Osthus, PhD

Named author institution: Los Alamos National Laboratory, Statistical Sciences Group Funding: This work was supported by LANL-LDRD ER grant 20200700ER

Other team authors:

Los Alamos National Laboratory, Information Systems and Modeling: Sara Del Valle, Carrie Manore, Lauren Castro, Courtney Shelley, Mandy Smith, Julie Spencer, Geoffrey Fairchild, Dax Gerts, Chrysm Watson Ross, Lori Dauelsberg, Ashlynn Daughton, Morgan Gorris, Beth Hornbein, Nidhi Parikh, Deborah Shutt

Los Alamos National Laboratory, Intelligence and Systems Analysis: Travis Pitts

Los Alamos National Laboratory, Statistical Sciences Group: Isaac Michaud, Brian Weaver Los Alamos National Laboratory, Space Data Science and Systems: Amanda Ziemann

Los Alamos National Laboratory, Continuum Models and Numerical Methods: Daniel Israel

Team name: Oliver Wyman – Navigator

Named Author: Ugur Koyluoglu, PhD

Named author institution: Oliver Wyman

Other team authors:

Oliver Wyman - New York: Gokce Ozcan, John Milliken, James Morgan, Michael Moloney, Helen Leis, Bruce Hamory, M.D., Chris Schrader, Christina Kyriakides, Daniel Siegel, Alexander Wong, David DesRoches

Oliver Wyman - Toronto: Chris Steifeling

Oliver Wyman - London: Barrie Wilkinson

Team name: Columbia University

Named Author: Teresa K Yamana, PhD

Named author institution: Columbia University, Mailman School of Public Health, Dept. of Environmental Health Sciences

Other team authors:

Columbia University, Mailman School of Public Health, Dept. of Environmental Health Sciences: Sen Pei, Marta Galanti, Jeffrey Shaman

Columbia University, Mailman School of Public Health, Dept. of Epidemiology: Wan Yang

Team name: COVID-19 Policy Alliance

Named Author: Andrew Zheng, M.S.

Named author institution: Operations Research Center, Massachusetts Institute of Technology

Other team authors:

Operations Research Center, MIT: Jackie Baek, Andreea Georgescu, Joshua Wilde, Deeksha Sinha

Sloan School of Management, MIT: Retsef Levi, Vivek Farias

Team name: Epiforecasts

Named Author: Katharine Sherratt, MSc

Named author institution: London School of Hygiene and Tropical Medicine

Other team authors:

London School of Hygiene and Tropical Medicine: Nikos I Bosse, Sophie Meakin, Sam Abbott, Joel Hellewell, James D Munday, Sebastian Funk

Funding: This work was supported by the Wellcome Trust [210758]

Team name: COVID-19 Simulator

Named Author: Jagpreet Chhatwal, PhD

Named author institution: Massachusetts General Hospital, Harvard Medical School Other team authors:

Massachusetts General Hospital Institute for Technology Assessment: Peter Mueller, Madeline Adee, Mary Ann Ladd

Georgia Institute of Technology, H. Milton Stewart School of Industrial and Systems Engineering: Turgay Ayer, Yingying Xiao

Value Analytics Labs: Ozden O Dalgic

Boston University School of Medicine, Boston Medical Center: Benjamin P Linas Funding: National Science Foundation awards 2035360 and 2035361

Team name: Iowa State - Lily Wang’s Research Group

Named Author: Lily Wang, Ph.D.

Named author institution: Department of Statistics, Iowa State University

Other team authors:

College of William & Mary, Department of Mathematics: Guannan Wang

Iowa State University, Department of Finance: Lei Gao

Statistical and Applied Mathematical Sciences Institute / University of North Carolina at Chapel Hill: Xinyi Li

Iowa State University, Department of Statistics: Shan Yu, Myungjin Kim, Yueying Wang, Zhiling Gu

Funding and acknowledgments: Zhiling Gu was supported in part by National Science Foundation award DMS-1916204. Myungjin Kim was supported in part by National Science

Foundation award DMS-1934884. The computing was supported by College of Liberal Arts and Sciences, Iowa State University.

Team name: NotreDame-FRED

Named Author: Guido España, PhD

Named author institution: University of Notre Dame

Other team authors:

University of Notre Dame: Rachel Oidtman, Sean Cavany, Alan Costello, Annaliese Wieler, Anita Lerch, Carly Barbera, Marya Poterek, Quan Tran, Sean Moore, and T. Alex Perkins Funding: NSF RAPID grant. Real-time updating of an agent-based model to inform COVID-19 mitigation strategies. Award number 2027718.

Team name: IHME COVID-19

Named Author: Robert C Reiner, Jr, PhD

Named author institution: University of Washington, School of Medicine, Department of Health Metrics Sciences, Institute of Health Metrics Sciences

Other team authors:

IHME: David Pigott, Ryan Barber, Chris Odell, Steve Lim, Simon Hay, Emmanuela Gakidou, Chris Murray

Funding: Bill & Melinda Gates Foundation and National Science Foundation award 2031096

Team name: NotreDame-mobility

Named Author: Sean Cavany, PhD

Named Author institution: University of Notre Dame, Department of Biological Sciences Other team authors:

University of Notre Dame: Rachel Oidtman, Sean Cavany, Alan Costello, Annaliese Wieler, Anita Lerch, Carly Barbera, Marya Poterek, Quan Tran, Sean Moore, and T. Alex Perkins Funding: GE and TAP were supported by a RAPID grant from the National Science Foundation (DEB 2027718).

Team name: USACE ERDC

Named Author: Glover E. George, PhD

Named author institution: US Army Engineer Research and Development Center Other team authors:

Information Technology Laboratory: Ian Dettwiller, William England, Glover George, Harrison Hunter

Environmental Laboratory: Brandon Lafferty, Micheal Mayo, Micheal Rowland

Cold Regions Research and Engineering Laboratory: Matthew Parno

Coastal Hydraulics Laboratory: Matthew Farthing

Funding: Funding provided by the US Army Corps of Engineers Geospatial Task Force.

Team name: MOBS-GLEAM

Named Author: Xinyue Xiong, BS

Named author institution: Northeastern University, Network Science Institute

Other team authors:

Northeastern University: Matteo Chinazzi, Jessica T. Davis, Kunpeng Mu, Ana Pastore y Piontti, Alessandro Vespignani

